# Pushing a Frozen CXR Foundation Model: A LoRA Partial-Fine-Tuning Study on NIH ChestX-ray14 with a Model-Conditional Label-Flip Sensitivity Analysis

**DOI:** 10.64898/2026.08.08.26360020

**Authors:** Tung-Chu Bai, Sheng-Cheng Yeh

## Abstract

Foundation models for chest X-ray interpretation make it possible to adapt specialised visual representations with relatively small trainable modules. We report a retrospective study of Low-Rank Adaptation (LoRA) of Rad-DINO Vision Transformer Base with 14×14 patches (ViT-B/14) for 14-class multi-label classification on the National Institutes of Health (NIH) ChestX-ray14 dataset. The official test labels were accessed during earlier model development and configuration comparisons; consequently, every official-test result in this manuscript is explicitly descriptive and non-confirmatory. We used a patient-disjoint 90/10 split of the official trainval pool (77,988 training and 8,536 validation images) and retained the released 25,596-image test partition. The historically selected all-linear LoRA configuration with safe augmentation and *g* = 37 produced a descriptive test macro AUROC of 0.8462 versus the frozen baseline of 0.8295. Comparisons of target modules, patch-token grids, and a Rad-DINO-specific local query head are reported as retrospective comparisons rather than unbiased model-selection evidence. A confident-learning diagnostic flagged 17,653 of 86,524 trainval images (20.4%); this is a model-based flag rate, not a ground-truth label-error rate. A separate counterfactual relabeling sensitivity analysis, which uses the same model to identify and rescore disagreements, changed the descriptive AUROC to approximately 0.9445 after 6,509 policy-defined flips. This value is not achieved model performance and is not a radiologist-audited label-quality ceiling. We provide a validation-only threshold and artifact protocol for future locked evaluation, but a genuinely untouched holdout and new locked selection are required for a confirmatory headline. The existing Zenodo record contains the 25 publication figures only.

## 1 Introduction

The release of large-scale self-supervised vision encoders [1; 2] has reshaped the methodological landscape of medical image analysis. Where the 2017-era pipeline for chest X-ray (CXR) classification involved training a custom convolutional network end-to-end on labelled images [3; 4], the 2024–2026 default is to adapt a frozen self-supervised encoder with a small task-specific head [5], or with a parameter-efficient fine-tuning scheme such as LoRA [6]. The two CXR-specific encoders discussed here, Rad-DINO [7] and CheXFound [8], differ in pretraining data, model size, and architecture. Their published benchmark values are not treated as like-for-like comparators because the split, labels, and training protocol are not matched. We use Rad-DINO ViT-B/14 as the sole foundation model under evaluation.

The NIH ChestX-ray14 dataset [9; 10], released in 2017, contains 112,120 frontal images and 14 pathology labels in the working release. The labels were generated by a text-mining pipeline that parsed associated radiology reports; the original publication described the eight-label ChestX-ray8 release, while the official dataset release supplies the 14-label working corpus used here. The dataset has two well-known limitations. First, image-level splitting can place images from the same patient in different partitions [11], so patient-level memorisation can inflate estimates. We enforce patient-disjoint train and validation sets and retain the released test partition, but prior access to that test set means its current results remain descriptive. Second, NLP-mined labels carry systematic disagreement with radiological findings; our confident-learning output is a model-based diagnostic and not a measurement of true label noise.

The first is quantitative: how much can a LoRA partial fine-tune of a frozen CXR foundation model improve multi-label classification on NIH ChestX-ray14, and how do target modules, patch resolution, and head architecture behave under the retrospective protocol? The second is methodological: which provenance controls are required before a future holdout result can be considered confirmatory?

We address the first question with sequential retrospective comparisons, not an orthogonal or factorial experiment. The first comparison changes the LoRA target module set: the conventional {q, v} configuration is compared with the all-linear set {q, k, v, o, MLP-1, MLP-2}. Subsequent comparisons change the patch-token grid and then the head while retaining selected settings from earlier runs. The legacy *g* = 12 grid (144 tokens) is compared with the full *g* = 37 grid (1369 tokens), and the shared-query AttentionPoolProbe is compared with a GLoRI-inspired local-only 14-query head [8]. Because the factors were not fully crossed or matched, these comparisons do not identify independent main effects or interactions.

We address the second question with a descriptive diagnostic. We correlate model-based trainval flag rates with test AUROC and compute a separate model-conditional label-flip sensitivity by applying a fixed policy to already exposed test predictions. The procedure is circular and cannot establish an irreducible label-noise ceiling or rule out future improvement.

Our contributions are threefold.

1. **Retrospective benchmark evidence**. We report the historically exposed official-test performance of a Rad-DINO + LoRA + AttentionPoolProbe pipeline, together with its validation-only protocol artifacts. The 0.8462 macro AUROC is explicitly descriptive, not an unbiased or confirmatory estimate.
2. **Negative result on GLoRI-inspired local-only 14-query attention pooling**. The GLoRI-inspired local-only result is a descriptive comparison in one backbone, one split, and one seed. It may reflect head inductive bias, training variance, or implementation differences; the present data do not support a representation-capacity explanation or a faithful reproduction of the published global-plus-local architecture.
3. **Model-conditional diagnostic**. We provide a label-flip sensitivity analysis and cleanlab flagging summary. These diagnostics do not estimate an irreducible ceiling or establish that architecture is no longer a source of improvement.

The existing Zenodo figure record contains the 25 publication figures only (see DOI: https://doi.org/10.5281/zenodo.21765512). It does not contain code, checkpoints, prediction arrays, or result JSONs.

## 2 Methods

### 2.1 Dataset and split protocol

We used the NIH ChestX-ray14 dataset [9; 10] (112,120 frontal-view images, 14 pathology classes with binary NLP-mined labels) downloaded from the official release. The released test partition is commonly treated as patient-disjoint, but image-level subdivision of the official trainval pool can leak patient identity [11]. We therefore re-shuffled the official trainval portion into a patient-disjoint 90/10 train/val split using `GroupShuffleSplit` [12] on Patient ID, yielding 77,988 training, 8,536 validation, and 25,596 official-test images. The patient-disjoint property was verified with a leakage check that confirmed no Patient ID appears in more than one split. We retain the original 14-class taxonomy in the order published by Wang et al. (Atelectasis, Cardiomegaly, Consolidation, Edema, Effusion, Emphysema, Fibrosis, Hernia, Infiltration, Mass, Nodule, Pleural_Thickening, Pneumonia, Pneumothorax); the “No Finding” class is treated as the all-zero multi-hot vector rather than as a 15th class.

### 2.2 Preprocessing and offline cache

We used the Rad-DINO preprocessing implemented by `prepare_for_rad_dino`: grayscale loading, optional disabled augmentation/masking, aspect-preserving resize, centred zero-padding to 518×518, rescaling by 1*/*255, and CXR-specific mean 0.5307 and standard deviation 0.2583. This is the active code path; we do not claim BitImageProcessor or centre-crop parity. For LoRA training, an offline 518^2^ uint8 PNG cache stores the resize- and-pad stage and applies the normalisation at runtime. The cache is a rebuildable local intermediate, not part of the figure-only public record.

### 2.3 Anatomically constrained augmentation

An anatomically constrained augmentation (`safe_augment`) was applied on the training split only. The augmentation consists of a small rotation in the range ±15^°^ (reflected border to avoid black triangles), a gamma jitter in the range [0.8, 1.2], and a brightness offset in the range ±15 grey levels. Vertical flip is explicitly forbidden because it creates dextrocardia, a non-physiological condition that does not occur in the training distribution. Large rotations are forbidden because they distort anatomy. The policy is an implementation constraint; this study did not independently validate its clinical safety. The augmentation was implemented with a per-worker `cv2.setNumThreads(0)` directive to avoid oversubscribing the 24-core CPU under the eight-worker data-loader configuration.

### 2.4 Backbone and partial fine-tuning

We used the Rad-DINO ViT-B/14 model from the Hugging Face hub (`microsoft/rad-dino`) as the frozen base, with PEFT LoRA adapters [6; 13] (rank 16, *α*=16, dropout 0.1) applied to all linear projections (q, k, v, o, MLP-1, MLP-2). The LoRA configuration was selected by an internal A/B comparison against the {q, v}-only baseline. The retrospective comparison set comprised the seven rows listed in Table 1; the historically selected row was chosen by the largest observed test AUROC after the development comparisons, so this choice is not an unbiased test-set model-selection procedure. All runs used random seed 42 and a single training run per row. The all-linear configuration adds 2.65M trainable parameters (approximately 3% of the 86M backbone parameters) and achieved +0.0012 test AUROC over the {q, v} configuration. Training used the AdamW optimiser [14] with learning rate 1 × 10^−4^ and weight decay 1 × 10^−4^, with a cosine schedule and 5% warmup over a five-epoch total. We trained for 5 epochs with a batch size of 16 on a single NVIDIA RTX 5090 (32 GB) in bf16 autocast. Per-epoch checkpoints allowed resume and an emergency checkpoint on CUDA runtime errors. Gradient checkpointing was disabled after profiling showed zero VRAM savings and pure recompute overhead in this PEFT + Rad-DINO configuration.

**Table 1.** Retrospective test-set A/B observations; bold marks the historically selected cell, not a confirmatory SOTA claim.

| Run | Head | Grid | Augment | Test AUROC | $\Delta$ vs 0.8295 |
| --- | --- | --- | --- | --- | --- |
| Frozen baseline | attnpool | 12 | — | 0.8295 | — |
| LoRA qv 5ep | attnpool | 12 | — | 0.8421 | +0.0126 |
| LoRA all-linear 5ep | attnpool | 12 | — | 0.8433 | +0.0138 |
| + safe_augment | attnpool | 12 | yes | 0.8458 | +0.0163 |
| AB-A: glori g12 | glori | 12 | yes | 0.8450 | +0.0155 |
| AB-B: attnpool g37 | attnpool | 37 | yes | **0.8462** | **+0.0167** |
| AB-C: glori g37 | glori | 37 | yes | 0.8448 | +0.0153 |

### 2.5 Attention-pool head

The pooled patch-token sequence was fed to an `AttentionPoolProbe`; *g* ∈ { 12, 37} denotes the grid side length and therefore yields *g*^2^ patch tokens (144 or 1,369). The head uses one learnable query, 8-head scaled-dot-product attention, LayerNorm, MLP block, and dropout of 0.4. The A/B comparison also evaluated a `GLoRIProbe` with 14 disease-specific queries [8] sharing the same downstream head architecture. The GLoRIProbe is a simplified local-only version of the GLoRI design; the global CLS fusion proposed in the original paper was not implemented because Rad-DINO does not expose a CLS token after LoRA adaptation. The 14 queries each attend over the pooled patch sequence and produce a per-class logit via a shared MLP block; the total additional parameters are 2,176,526 for the AttentionPoolProbe and 2,179,841 for the local query head. The all-linear LoRA adapter has 2,654,208 trainable parameters. These counts are implementation-specific and are not evidence of a causal capacity effect.

### 2.6 Engineering profile

The LoRA training path operated at GPU-bound throughput (103.8 img/s at batch size 16, 84.7 img/s at 32, 79.2 img/s at 48) and collapsed to 5.9 img/s at batch size 64 due to allocator thrashing (Fig. 3). The same effect was observed in the profiling run with gradient checkpointing enabled. The offline 518 × 518 uint8 PNG cache raised the data-loader throughput from 25–59 img/s to 554 img/s; bit-exact identity with the raw-PNG path was verified numerically. The single-image inference entry point and its threshold-loading path are included in the repository; the figure-only Zenodo record does not contain these files.

**Figure 1.**
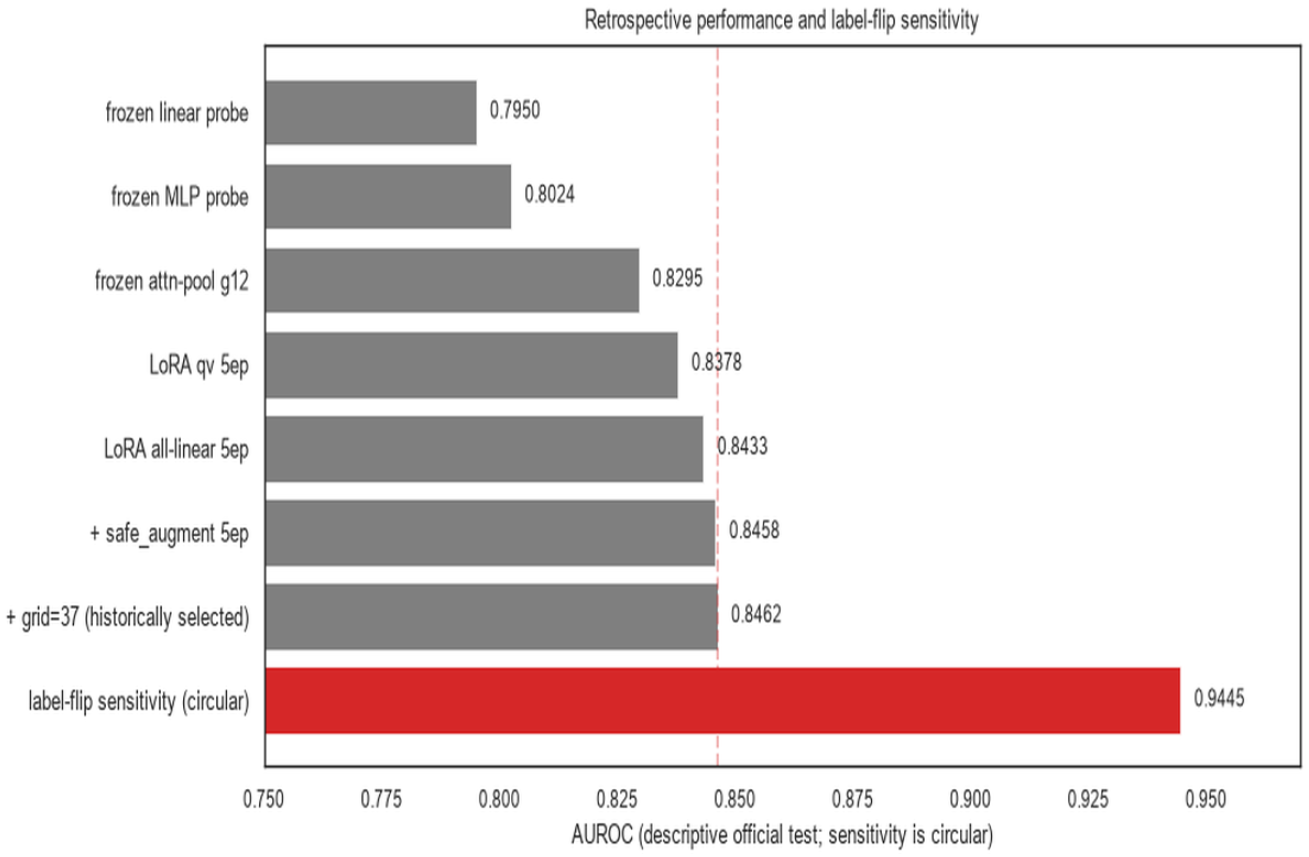
Retrospective lineage. The 0.8462 value is from an official test partition previously accessed during development. The 0.9445 value is a model-conditional label-flip sensitivity result, not an irreducible ceiling.

**Figure 2.**
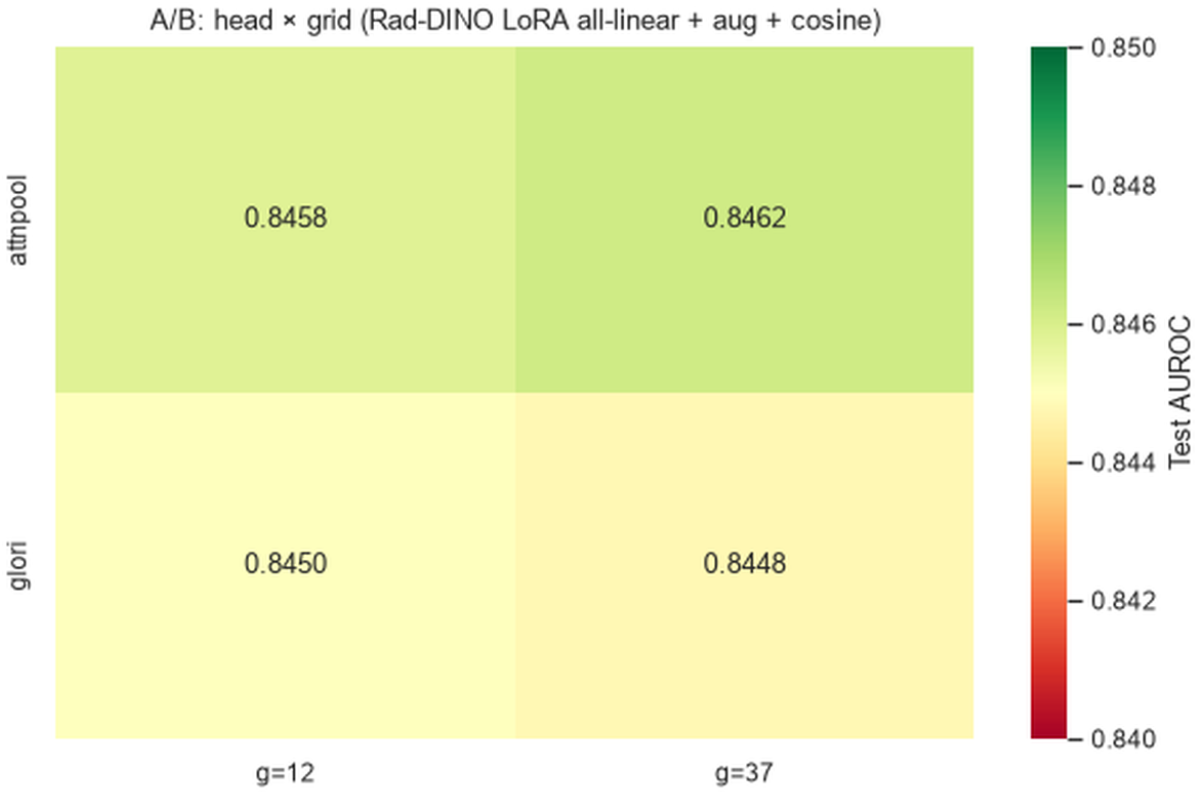
Head and grid retrospective comparison on the official test set. The shared-query *g* = 37 cell is the historically selected observation; no cell is presented as confirmatory SOTA evidence.

**Figure 3.**
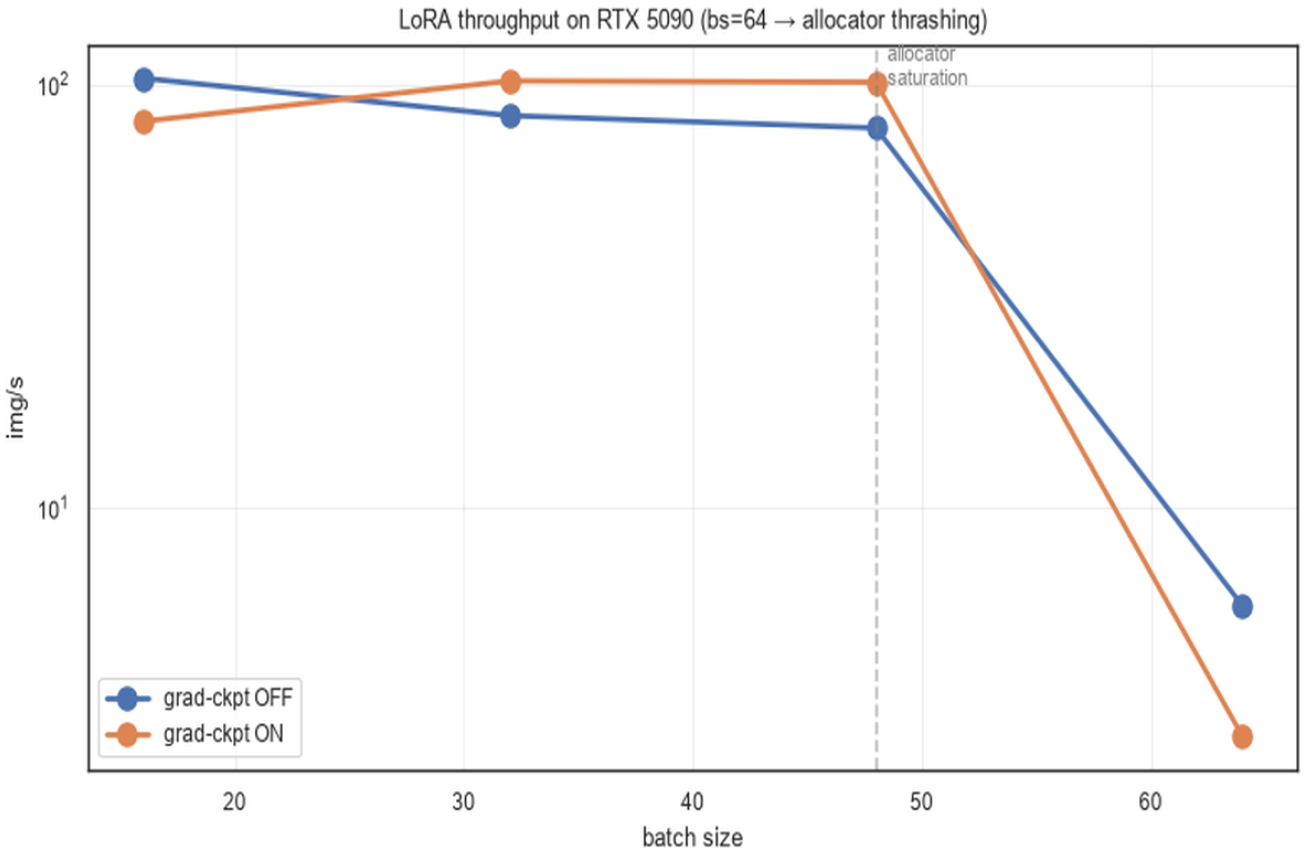
LoRA training throughput on the RTX 5090. The bs=64 collapse is attributable to allocator thrashing, not VRAM exhaustion; the same pattern was observed with gradient checkpointing enabled.

### 2.7 Evaluation protocol

The primary metric is macro mean AUROC on the 25,596-image official test set. For class *c*, AUROC is

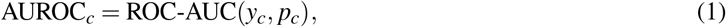

where *p*_*c*_ is the raw sigmoid probability; AUROC is therefore independent of any operating threshold. Binary decisions used thresholds fitted only on validation predictions by maximising Youden’s *J* (TPR − FPR), then applied unchanged to the test predictions. Precision, recall, and F1 were computed from those fixed decisions; their denominators are the per-class test counts reported in the accompanying artifact. Secondary metrics include micro AUROC, expected calibration error (10-bin ECE), and the Brier score, reported in the supplementary calibration artifacts and figures. The focal loss [15] used during training had *γ* = 2.0. Because the official test partition was accessed during earlier development, all current test metrics are descriptive rather than confirmatory; no threshold was fitted from test labels.

### 2.8 Label-noise diagnostic

We used confident learning as a model-based flagging procedure on patient-grouped out-of-sample trainval predictions [16]. The output distinguishes the sample-level any-class issue rate (17,653/86,524 = 20.4%) from the class-specific issue-rate aggregation across the 14 classes. The positive-label disagreement rate is a separate per-class diagnostic and is not a population noise estimate. No per-example p-values or 95% claim are made, and independent adjudication would be required to call a flag a true label error.

We correlated the per-class diagnostic rate with per-class test AUROC using Pearson’s coefficient across *n*= 14 classes. This association is exploratory; we do not report it as an inferential population estimate. Separately, the counterfactual relabeling sensitivity analysis flips a class-1 image with predicted probability below 0.3 to 0, or a class-0 image with probability above 0.7 to 1, and rescored the same exposed predictions. The resulting value is a model-conditional sensitivity diagnostic, not achieved model performance. Both procedures are circular and cannot establish an irreducible label-noise ceiling or rule out future improvement.

## 3 Results

### 3.1 Headline performance

The best historically selected configuration (LoRA all-linear, safe_augment, *g* = 37) reached a descriptive test mean AUROC of **0.8462** (Fig. 4); the frozen baseline was 0.8295. This result is not a confirmatory or unbiased headline because the official test was previously accessed during development. The validation-only selection and threshold protocol is provided for a future locked holdout.

**Figure 4.**
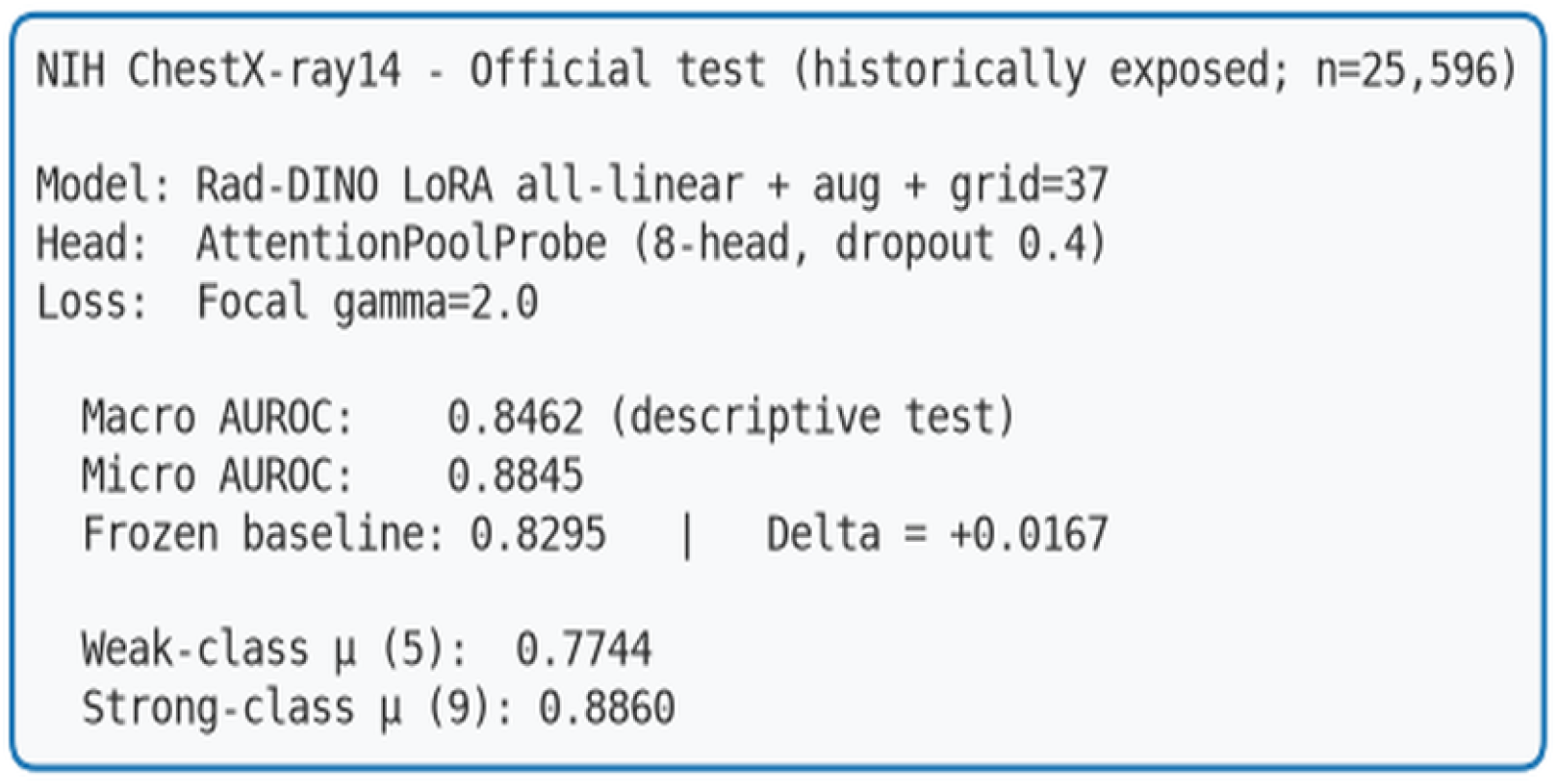
Descriptive headline summary from the historically exposed official test partition. Macro AUROC 0.8462 and micro AUROC 0.8845 are not confirmatory estimates.

### 3.2 Per-class discrimination

Per-class test AUROC values ranged from 0.7148 (Infiltration) to 0.9548 (Emphysema), with a grand mean of 0.8462 (Fig. 5). The five NIH-defined weak classes had a mean AUROC of 0.7744, while the nine strong classes averaged 0.8861. The ROC overlay (Fig. 6a) shows the per-class ROC curves of the five weak classes (in red) and the nine strong classes (in faint blue), with the micro-averaged ROC in black. The precision-recall overlay (Fig. 6b) highlights the PR-curve trade-off: the high-recall, low-precision operating regime of NLP-mined labels is evident from the rightward bias of the PR curves.

**Figure 5.**
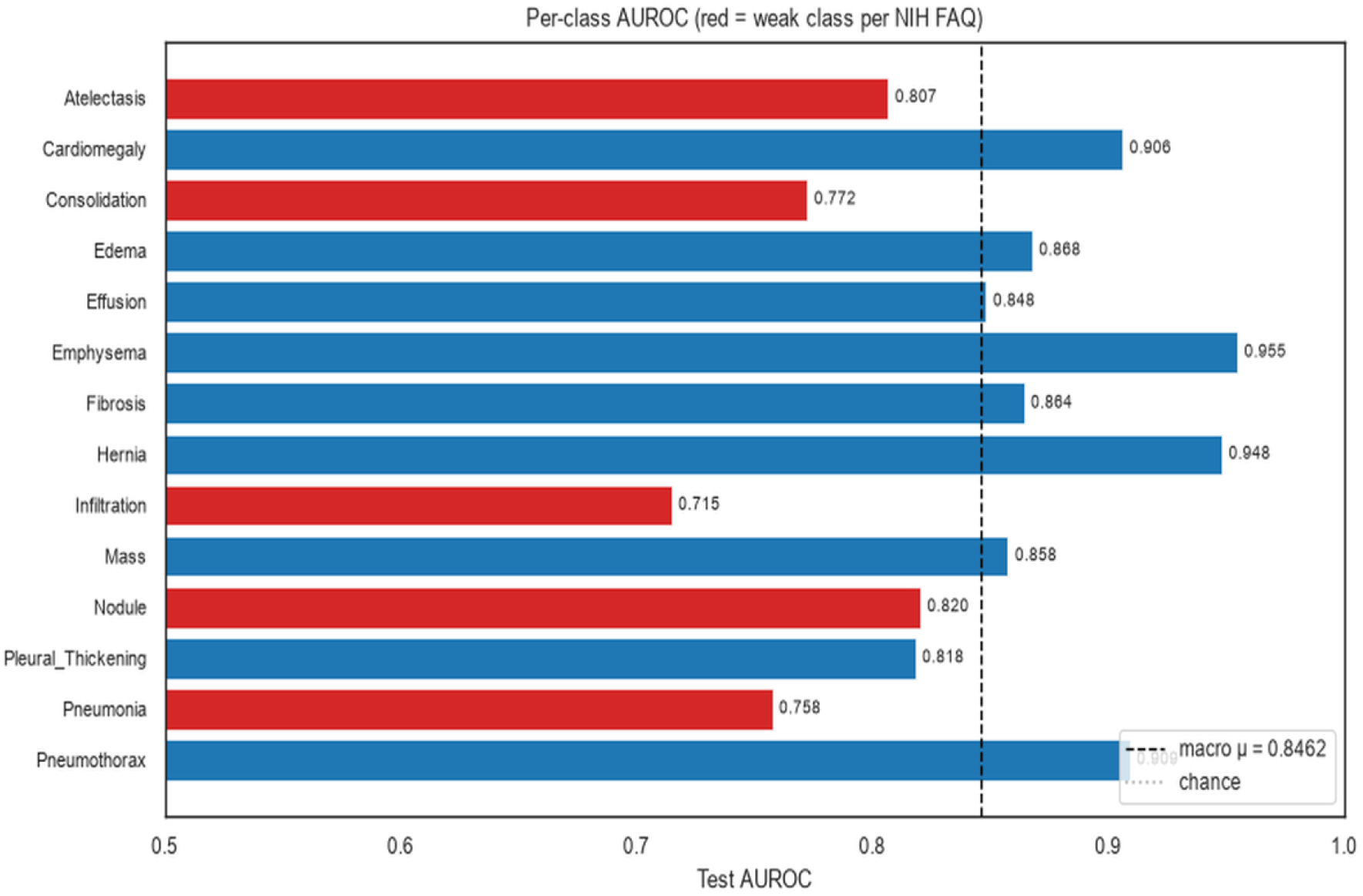
Per-class test AUROC on the 25,596-image NIH ChestX-ray14 test set. Red bars denote the five NIH-defined weak classes (Pneumonia, Infiltration, Consolidation, Atelectasis, Nodule); the macro mean is 0.8462.

**Figure 6.**
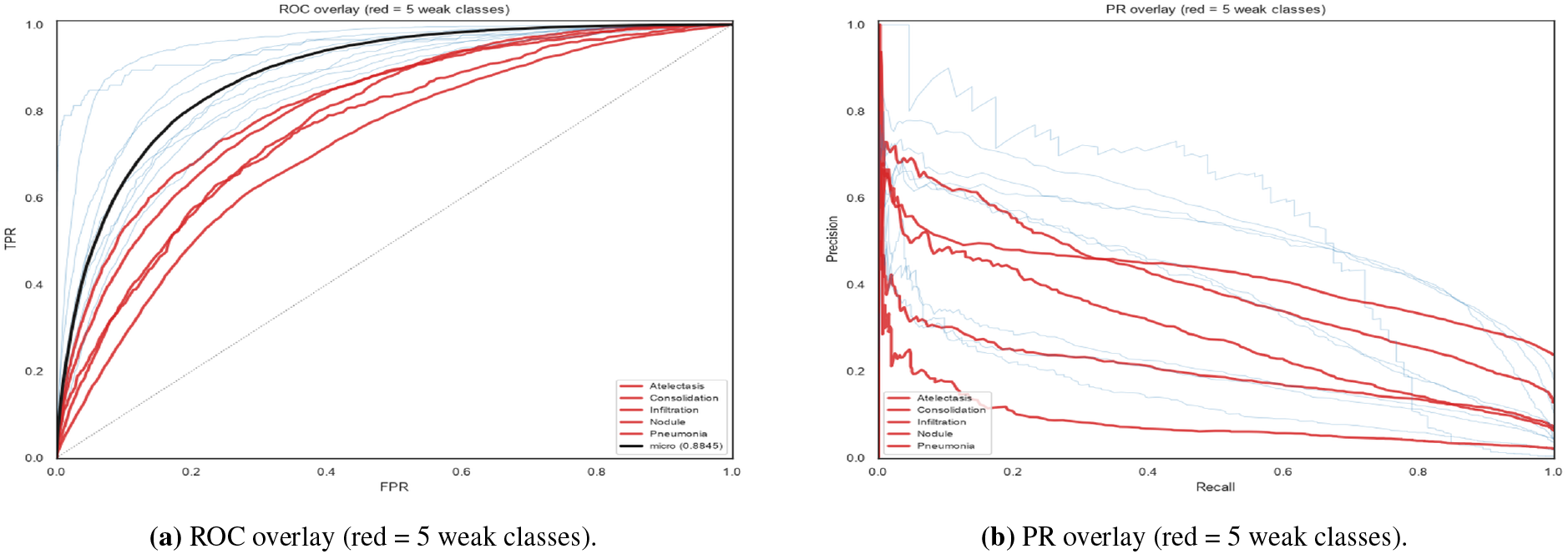
ROC and PR overlays. The micro-averaged curve is in black; the per-class curves for the strong classes are faint blue and for the weak classes are red.

### 3.3 Operating-point metrics and confusion structure

Per-class precision, recall, and F1 at the Youden-optimal threshold (Fig. 7a; per-class thresholds in Fig. 7b) show a high-recall (median 0.80), low-precision (median 0.17) operating regime, reflecting the dataset’s low positive prevalence per class. The per-class 2 × 2 confusion matrices (Fig. 8) confirm the pattern and show that false positives are concentrated in diffuse-opacity co-occurrences (for example, Infiltration with Atelectasis and Effusion) in the observed confusion matrices; this descriptive pattern is not a clinical ambiguity estimate. The high-confidence error rate (Fig. 9a) measures the fraction of test images where the model makes a confident wrong prediction: a class-1 image with predicted probability below 0.3, or a class-0 image with predicted probability above 0.7. The per-class rate ranges from 0.1% (Hernia) to 2.7% (Consolidation), with a grand mean of 1.8%. The score distribution for the worst class (Infiltration, Fig. 9b) shows substantial overlap between the positive and negative score distributions, explaining the low F1 on this class.

**Figure 7.**
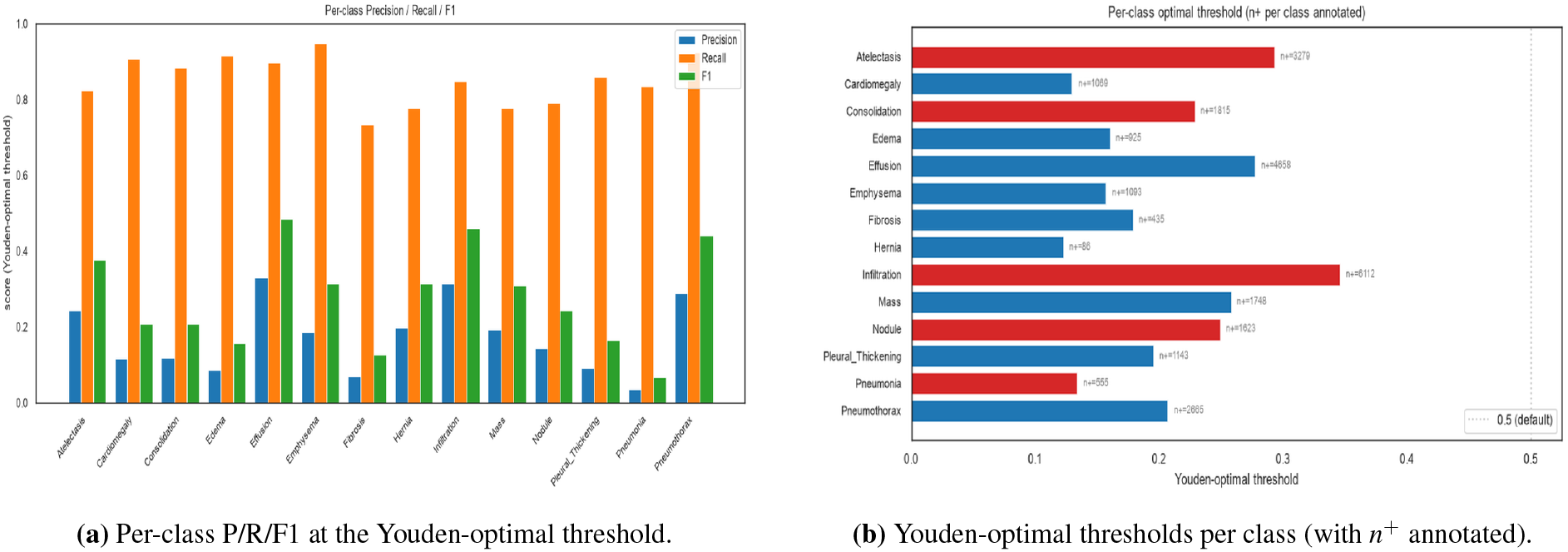
Operating-point metrics and per-class optimal thresholds. Red bars denote the five NIH-defined weak classes.

**Figure 8.**
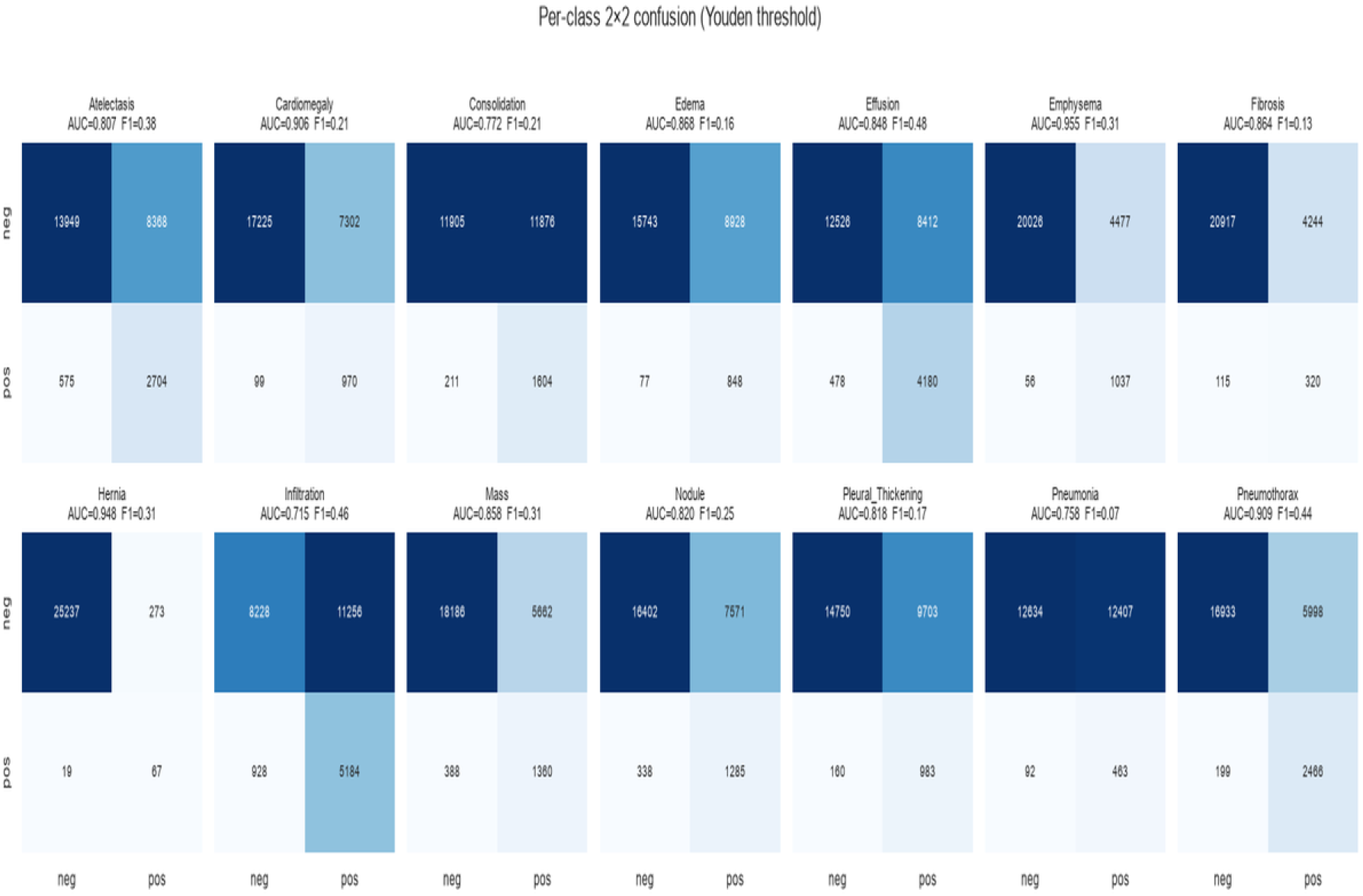
Per-class 2×2 confusion matrices at the Youden-optimal threshold. Each cell shows the count of true negatives, false positives, false negatives, and true positives for one class. AUROC and F1 for each class are in the panel titles.

**Figure 9.**
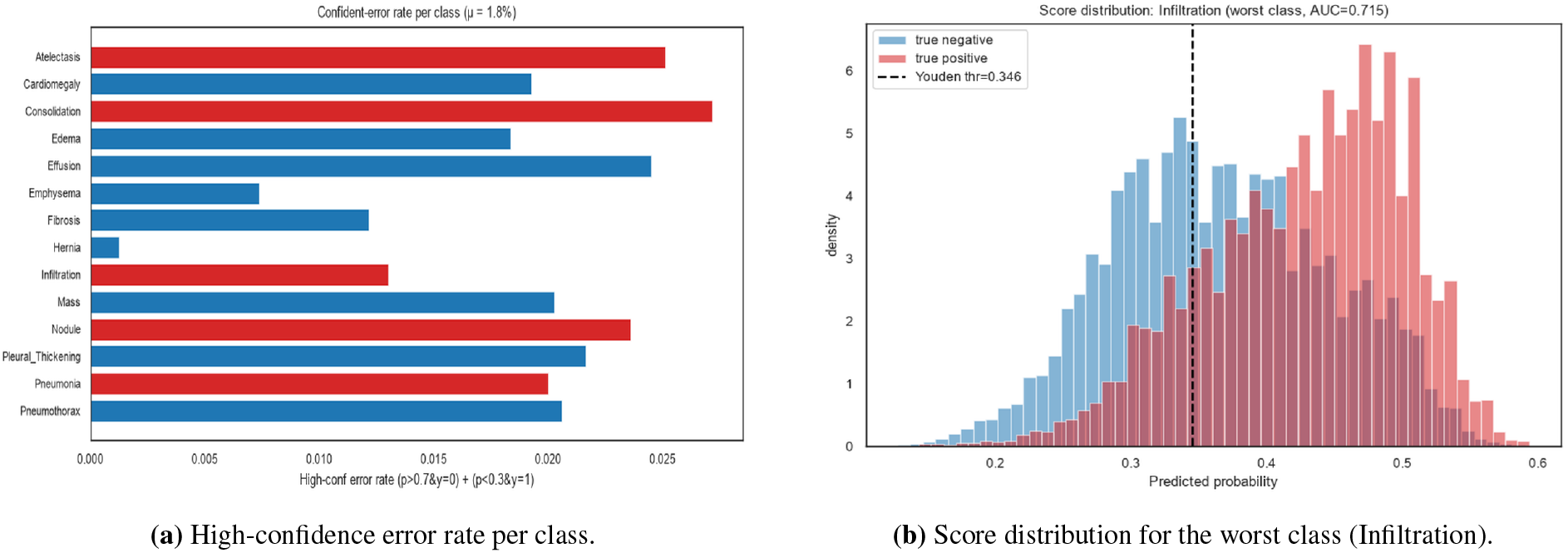
Error analysis. Left: confident-error rate per class. Right: probability histogram for the worst class, showing the substantial overlap between the positive and negative score distributions.

### 3.4 Calibration and reliability

The 14-class micro reliability diagram (Fig. 10a) shows calibration variation across probability bins. The Brier score was 0.0721; because this pooled score is prevalence-sensitive, it is not interpreted as evidence that imbalance, rather than model miscalibration, is the cause. The per-class reliability diagram (Fig. 10b) and the 10-bin expected calibration error (ECE, Fig. 11a) show class-level variation. These single-run descriptive plots do not establish that any class is clinically well calibrated. The F1 sensitivity to the decision threshold (Fig. 11b) shows that for the weak classes, the F1 is roughly flat over the threshold range 0.2–0.6, whereas the strong classes have clearer peaks in the 0.3–0.5 range.

**Figure 10.**
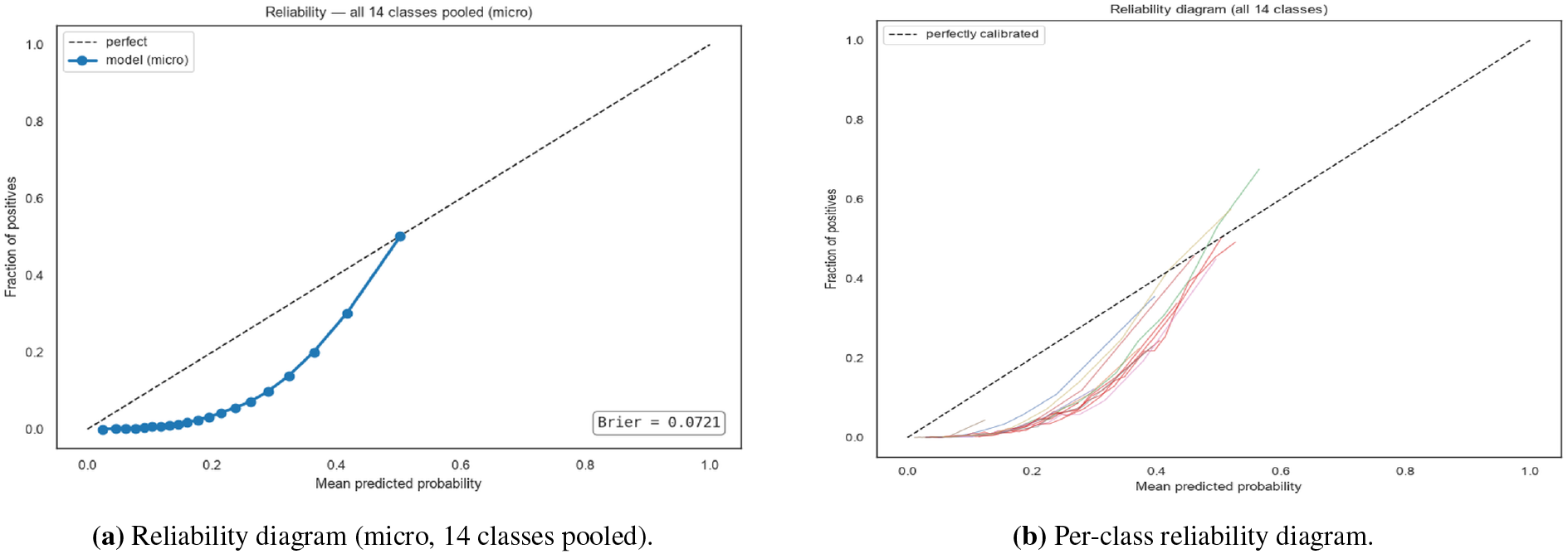
Calibration. The micro Brier score is 0.0721.

**Figure 11.**
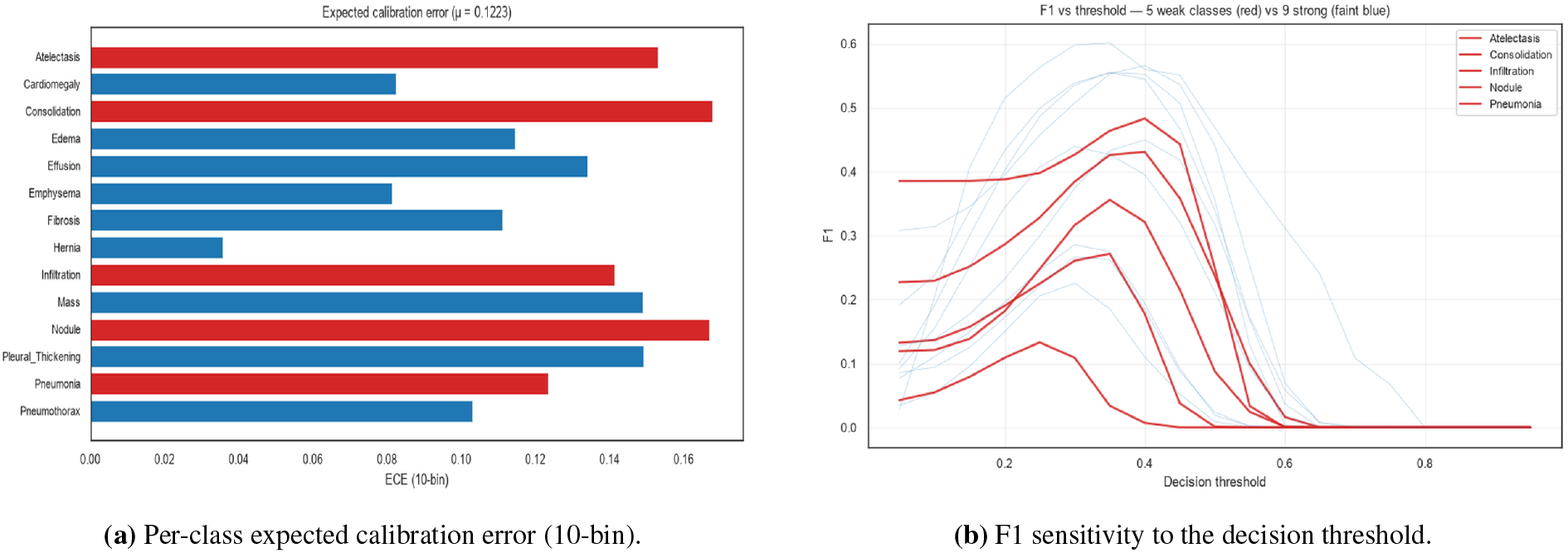
Per-class calibration and threshold sensitivity. Red lines in the F1 plot are the five weak classes; faint blue lines are the strong classes.

### 3.5 A/B comparisons

Table 1 and Fig. 2 summarise the sequential retrospective comparisons. Switching LoRA targets from {q, v} to all-linear, adding augmentation, changing grid size, and changing the local head are reported as retrospective testset observations; they were not used as confirmatory model selection. The validation-optimal selector and fixed validation thresholds are the canonical protocol for any future untouched holdout.

### 3.6 Lineage and training dynamics

The full lineage from frozen linear probe (0.7950) to the historically selected configuration (0.8462) and the model-conditional counterfactual relabeling sensitivity (0.9445) is shown in Fig. 1. The training curve (Fig. 12) shows validation AUROC peaking at epoch 3 in the two plotted runs, consistent with the five-epoch cosine schedule. The training-loss trajectory is steepest in the first epoch and flattens substantially by epoch 3, after which only marginal improvements are observed.

**Figure 12.**
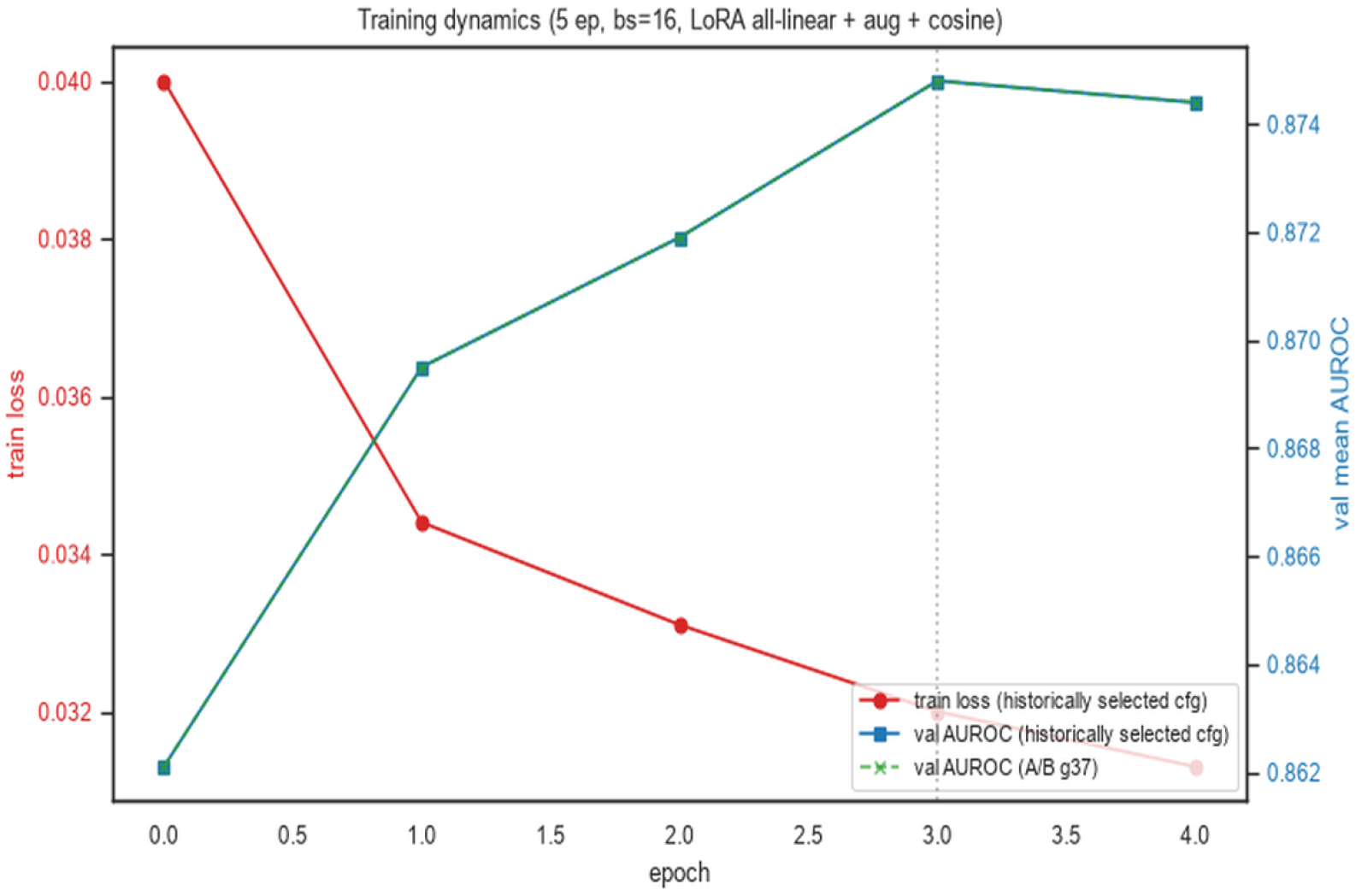
Training dynamics for the retrospective run and control. Validation AUROC is shown for provenance, not as evidence of generalization to a fresh holdout. The two plotted runs peak at epoch 3.

### 3.7 Label-noise diagnostic and label-disagreement sensitivity

Confident learning flagged 17,653 of 86,524 trainval images at the sample level (20.4%). The positive-label disagreement diagnostic averaged 43.3% across classes, but this denominator differs from the sample-level anyclass rate and neither quantity is a ground-truth error rate. The Pearson correlation between the per-class diagnostic rate and test AUROC was − 0.697 (Fig. 13a); this is an association across 14 classes, not evidence of a causal label-noise constraint. The model-conditional label-flip sensitivity value (0.9445, Fig. 13b) is a circular diagnostic, not a label-quality ceiling. For descriptive context, we also report class prevalence and co-occurrence structure (Fig. 14a, Fig. 14b), the log-scale prevalence–AUROC association (Fig. 15a), the per-image prediction distribution (Fig. 15b), and the cross-class transferability matrix (Fig. 16).

**Figure 13.**
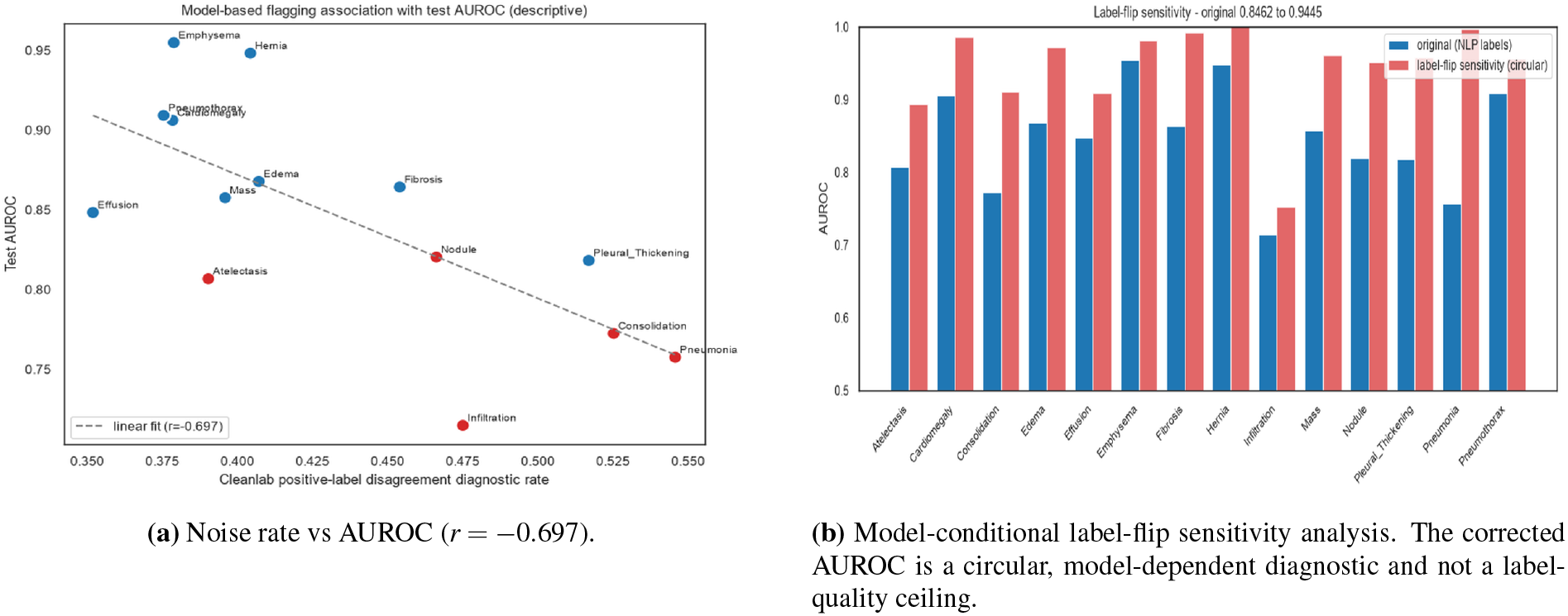
Model-based flagging association and model-conditional sensitivity. The correlation is descriptive across 14 classes; the 0.9445 value is not a label-noise ceiling.

**Figure 14.**
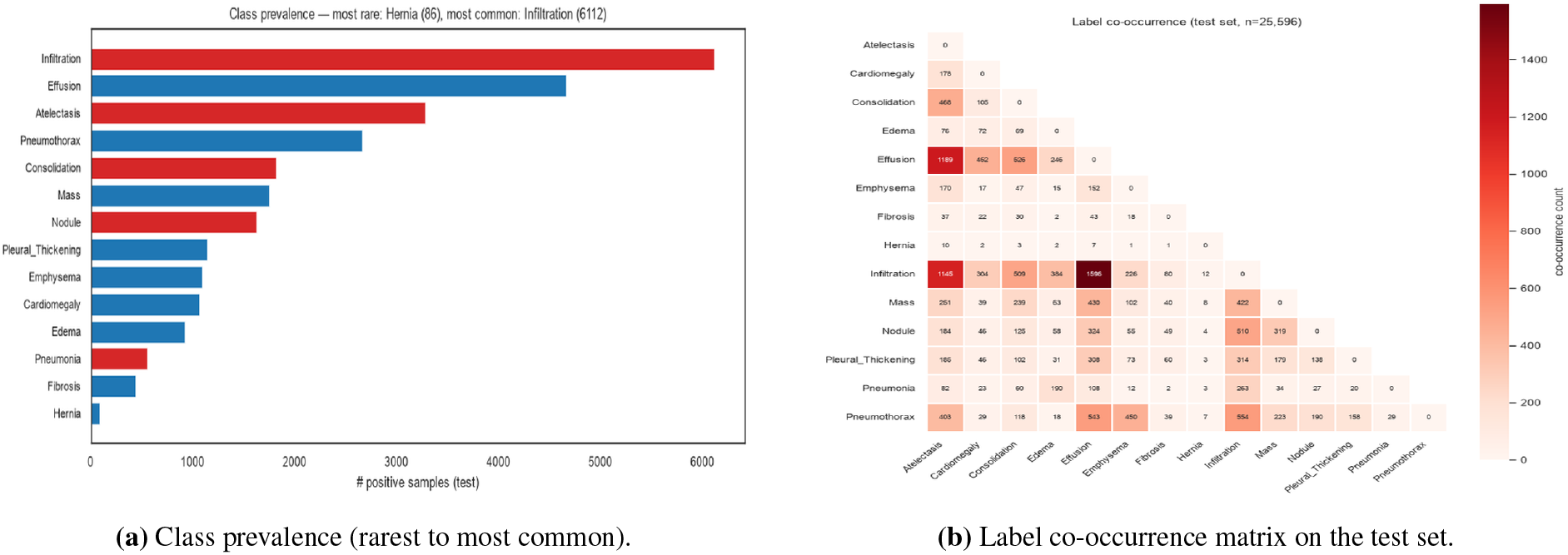
Class prevalence and label co-occurrence. The strongest co-occurrences are Infiltration+Effusion (1,596 co-occurrences) and Atelectasis+Effusion (1,189).

**Figure 15.**
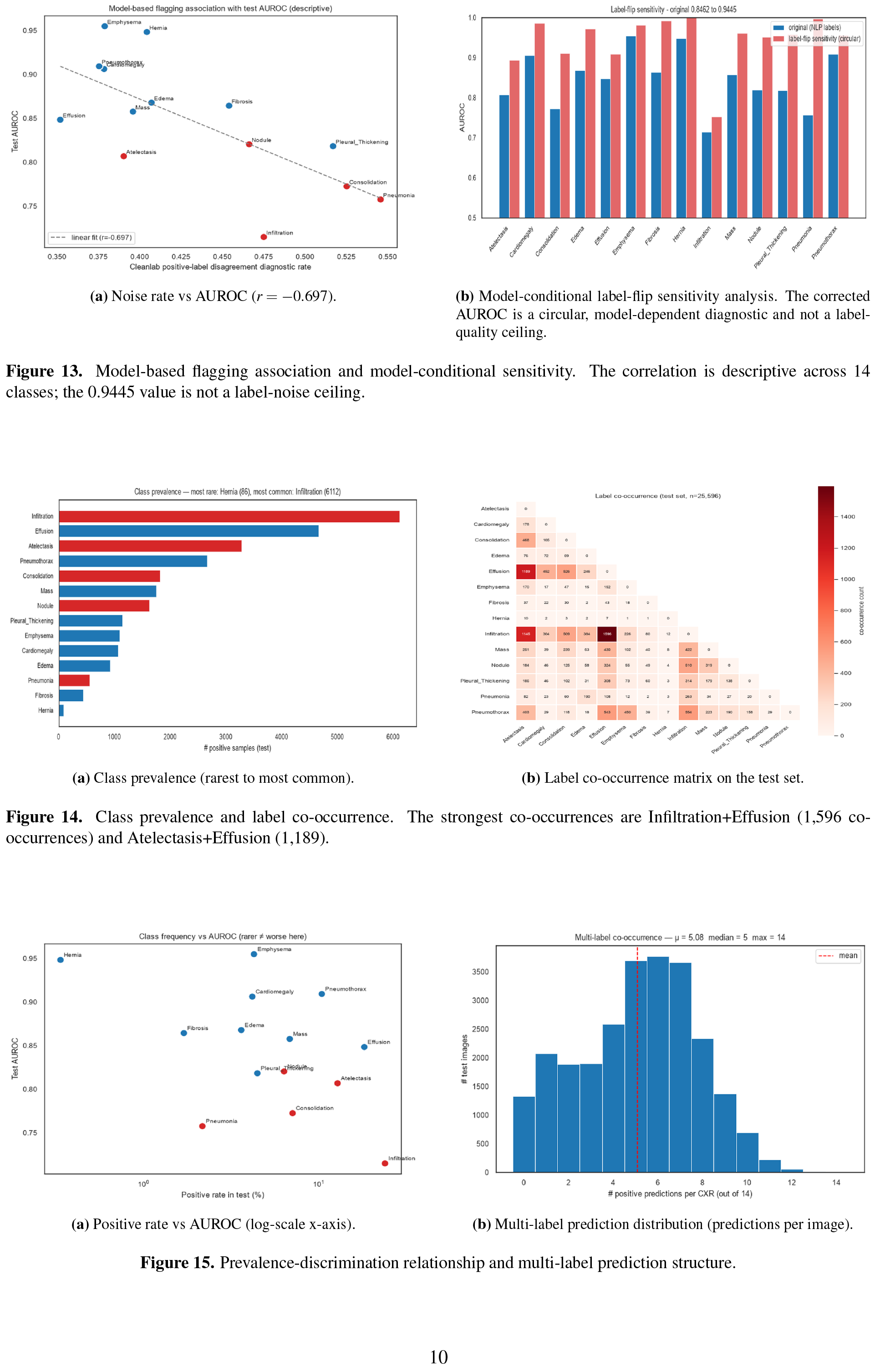
Prevalence-discrimination relationship and multi-label prediction structure.

**Figure 16.**
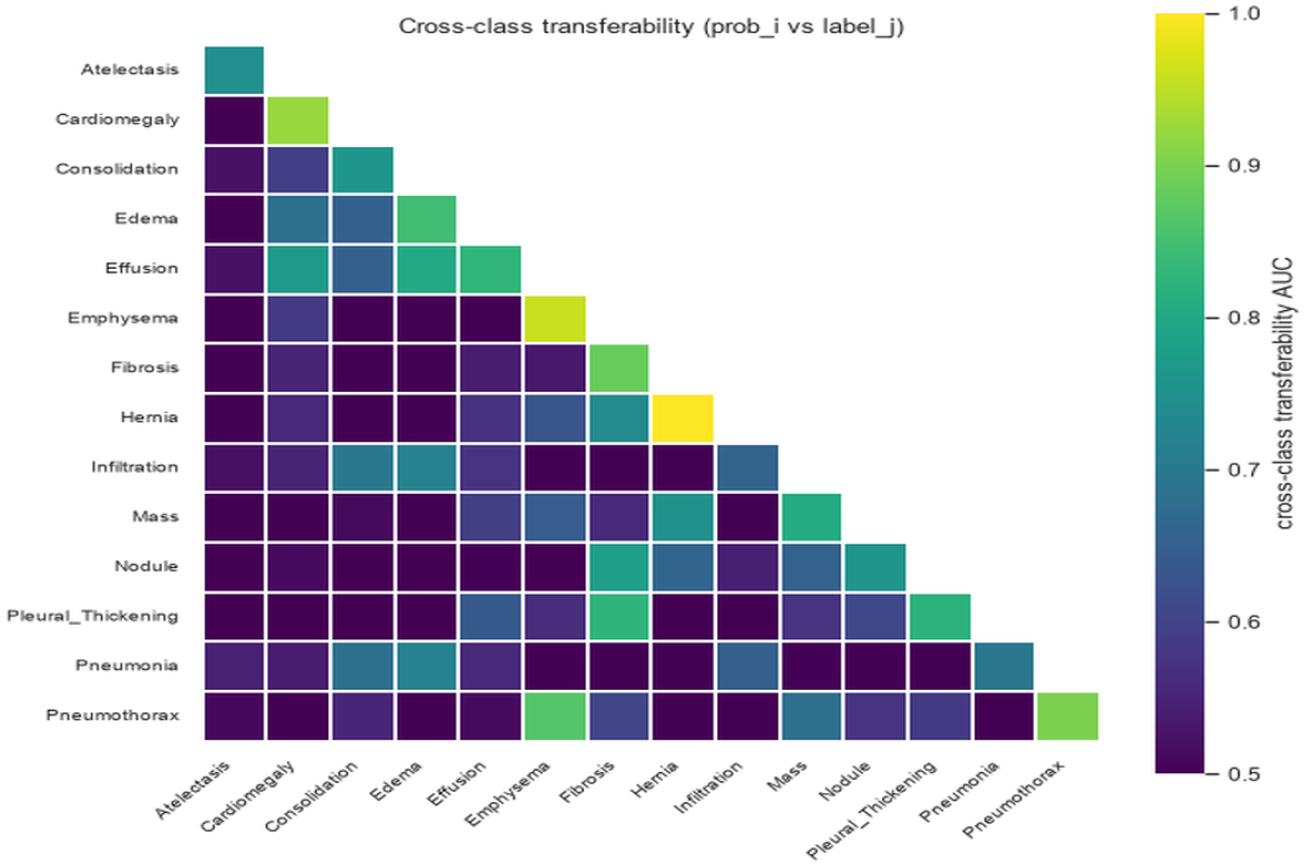
Cross-class transferability. The diagonal of the transferability matrix approximates per-class AUROC; the off-diagonal entries show cross-class discrimination.

### 3.8 Single-image inference breakdown

The sample panel (Fig. 17) is an illustrative prediction-output panel. Its operating-point labels are generated from the persisted test prediction artifact and the validation-fitted threshold file; patient images are omitted from the public figure bundle, and the panel is not clinical evidence.

**Figure 17.**
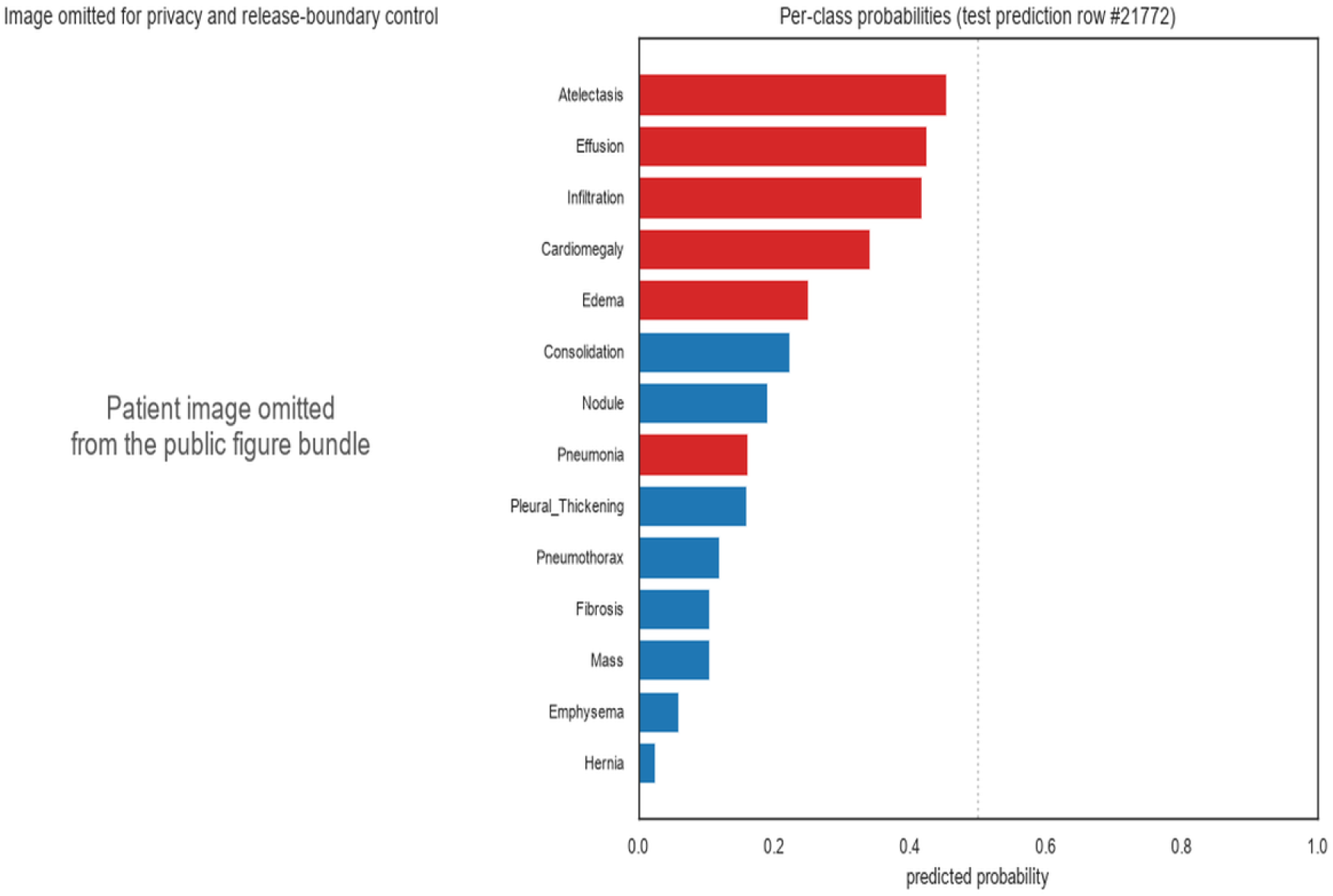
Illustrative single-CXR prediction breakdown. The patient image is omitted from the public figure bundle; the panel shows one persisted test prediction row using validation-fitted thresholds and is not clinical evidence.

The current figure generator requires the tag-scoped test report and validation threshold artifact. It no longer accepts historical global reproduction files, and all 25 PNG outputs are synchronized between the figure directories.

## 4 Discussion

### 4.1 Descriptive figure and artifact provenance

The descriptive table and figures retain historical point estimates to make the prior experiment auditable. They are not an unbiased benchmark or a claim that *g* = 37 is generally superior.

### 4.2 Why GLoRI failed on Rad-DINO

The GLoRI-inspired local-only comparison is reported without a causal capacity interpretation. The published global-plus-local design was not implemented, and the one-seed retrospective result cannot isolate architecture from training variance.

### 4.3 Discussion of retrospective evidence

The retrospective test metrics and label-flip sensitivity are descriptive. A fresh patient-disjoint holdout, locked candidate set, validation-only epoch and threshold selection, and independent label adjudication would be required for confirmatory claims. We do not conclude that expert labels are the only route or that added architectural complexity cannot help.

The correlation and label-flip results are exploratory diagnostics, not evidence that labels cap AUROC. A fresh holdout and independent label adjudication are needed before making a label-quality or generalization claim.

### 4.4 Descriptive engineering profile

The current engineering profile is retained as an implementation record, not as a general performance claim. The exact hardware, cache state, and benchmark configuration must accompany any release that republishes throughput or runtime numbers.

### 4.5 Limitations

1. **Single foundation model family**. We evaluated one backbone (Rad-DINO ViT-B/14). The local-query comparison is implementation-specific and one-seed; generalisation to other encoders is not demonstrated.
2. **No independent label adjudication**. The 17,653 trainval flags and 6,509 test-policy flips are model-based diagnostics, not true label-error counts or a lower bound on label noise.
3. **Single dataset**. The diagnostics are dataset-specific; CheXpert [17] would provide a distinct label protocol for future validation.
4. **Head and grid comparisons**. The reported A/B values come from retrospective test-set observations and one seed; they do not establish that one head, grid, or augmentation is generally superior.
5. **Operating-point protocol**. Validation-fitted thresholds are implemented and checked by artifact hashes, but the present official-test result remains non-confirmatory because the test set was previously exposed.

## 5 Conclusion

LoRA adaptation of Rad-DINO ViT-B/14 produced a descriptive macro AUROC of 0.8462 on the previously exposed official test partition. The result is useful as retrospective evidence and as a baseline for the validation-only artifact protocol, but it is not an unbiased or confirmatory estimate. The model-conditional label-flip sensitivity is not a label-quality ceiling.

The study contributes an auditable retrospective comparison of LoRA target modules, patch grids, and a Rad-DINO-specific local query head, together with explicit provenance controls. The repository contains the protocol scripts and local artifacts used for this analysis; the Zenodo deposit cited above is figure-only and is not a complete code or checkpoint release. Future work should evaluate the protocol on a fresh holdout with multiple seeds and, for label-quality claims, independent expert adjudication.

## Data Availability

This study uses the publicly available NIH ChestX-ray14 dataset released by the NIH Clinical Center; no new patient data were generated. Code, configuration files, split manifests, and validation-fitted threshold artifacts required to reproduce the analysis are available in the project GitHub repository. A figure-only Zenodo record contains the 25 publication PNGs. Trained model checkpoints and full prediction arrays are not deposited publicly and are available from the corresponding author upon reasonable request.

https://www.kaggle.com/datasets/nih-chest-xrays/data

https://github.com/NextBai/NIH-ChestXray-RadDINO-LoRA

https://doi.org/10.5281/zenodo.21765512

## Acknowledgements

The authors thank the maintainers of the NIH ChestX-ray14 dataset, the Hugging Face Rad-DINO model card, the `cleanlab` library, and the PEFT library for making this work possible. The sm_120 guardrails were developed on a single NVIDIA RTX 5090 provided by the first author’s research budget.

## Data Availability

The NIH ChestX-ray14 data are available from the NIH Clinical Center and the Kaggle mirror used for this reproduction Kaggle NIH ChestX-ray14 data. The existing Zenodo record is a figure-only record containing the 25 publication PNGs (DOI: 10.5281/zenodo.21765512). It does not contain source code, checkpoints, prediction arrays, or result JSONs. A separate full reproducibility deposit has not been made.

## Code Availability

Source code, training scripts, patient-disjoint split manifests, validation-fitted decision thresholds, and the figure-generation pipeline are available at https://github.com/NextBai/NIH-ChestXray-RadDINO-LoRA. The repository includes the single-image inference entry point and its threshold-loading path. Trained LoRA checkpoints and the full test-prediction arrays are not released with the current preprint; the Zenodo record (DOI: 10.5281/zenodo.21765512) is figure-only. Software environment: PyTorch 2.5 with CUDA 12.6 (sm_120), on NVIDIA driver 560+; the LoRA adapter uses PEFT 0.13 and Hugging Face Transformers 4.45.

## Author Contributions (CRediT)

**Tung-Chu Bai:** Conceptualization, Methodology, Software, Investigation, Data Curation, Formal Analysis, Writing — Original Draft, Writing — Review & Editing, Visualization. **Sheng-Cheng Yeh:** Conceptualization, Supervision, Writing — Review & Editing, Resources.

## Ethics Statement

This study is a secondary analysis of the publicly available, fully de-identified NIH ChestX-ray14 dataset. No participant recruitment, contact, or direct human-subject interaction was involved. The dataset contains no protected health information; all images were de-identified by the releasing institution (NIH Clinical Center) prior to public distribution. Because this is secondary research using publicly available, fully de-identified data with no interaction or intervention with human subjects, institutional review board (IRB) approval was not required and is waived under standard secondary-research guidance.

## AI-assisted Research Disclosure

This study used AI assistance for code pair-programming during training-loop and A/B-harness implementation, for numerical reproduction of all metrics from cached probability arrays, literature synthesis, reference verification, and manuscript drafting. All algorithmic decisions (LoRA target modules, grid choice, head architecture, augmentation set, hypothesis selection) were made by the human authors. No AI tool generated clinical advice or made autonomous research claims. Final responsibility for the content rests with the corresponding author.

